# Distinct temporal trends in breast cancer incidence from 1997 to 2016 by molecular subtypes: A population-based study of Scottish cancer registry data

**DOI:** 10.1101/19011411

**Authors:** Ines Mesa-Eguiagaray, Sarah H Wild, Philip S. Rosenberg, Sheila M Bird, David H Brewster, Peter S Hall, David Cameron, David Morrison, Jonine D Figueroa

## Abstract

**Background:** Strategies for breast cancer prevention are informed by assessing whether incidence differs by tumour biology. We describe temporal trends of breast cancer incidence by molecular subtypes in Scotland.

**Methods:** Population-based cancer registry data on 72,217 women diagnosed with incident primary breast cancer from 1997 to 2016 were analysed. Age-standardised rates (ASR) and age-specific incidence were estimated by tumour subtype after imputing the 8% of missing oestrogen receptor (ER) status. Joinpoint regression and age- period- cohort models were used to assess whether significant differences were observed in incidence trends by ER status.

**Results:** ER positive tumour incidence steadily increased particularly for women of screening age 50 to 69 years from 1997 till around 2011 (1.6%/year, 95%CI: 1.2 to 2.1). ER negative incidence decreased among all ages at a consistent rate of −0.7%/year (95%CI: −1.5, 0) from around 2000-2016. Compared to the 1941-1959 central birth cohort, women born 1912-1940 had lower incidence rate ratios (IRR) for ER+ tumours and women born 1960- 1986 had higher IRR for ER- tumours.

**Conclusions:** We show evidence of aetiologic heterogeneity of breast cancer. Future incidence and survival reporting should be monitored by molecular subtypes.

## Background

Breast cancer incidence is rising and it is the most common cancer among women worldwide (1). It is well established that breast cancer is not a single disease but comprises multiple subtypes, with oestrogen receptor (ER) expression a key marker of prognostic and aetiologic significance (2). ER+ tumours, which are amenable to targeted anti-oestrogenic therapies such as tamoxifen and aromatase inhibitors, are the most common type of breast cancers accounting for 65-75% of breast cancer cases in developed populations (3). Progesterone receptor (PR) is also a common marker of hormone responsiveness that is highly correlated with ER. Tumour over-expression of the human epidermal growth factor receptor 2 (HER2) was discovered two decades ago. This laid the foundation for biological therapies, which were shown to be clinically effective in treating tumours expressing HER2 and has been widely available in the UK since 2006 (4). ER- tumours do not express any of the hormone receptors described above and are rarer, have an earlier age of onset and worse prognosis than ER+ tumours, in part because fewer targeted treatments are available than for ER+ tumours. In addition to prognostic differences, epidemiologic studies have shown aetiologic differences by tumour subtypes (5, 6).

There are limited population cancer registries that collect ER, PR and HER2 data, the key distinguishing markers for molecular subtypes of breast cancer. Recent analyses support divergent incidence trends by ER status in the United States, Denmark and Ireland, with ER+ breast cancer incidence increasing and ER-breast cancer incidence decreasing (7-9). Data on combination of subtypes using ER, PR, and HER2 are even more limited, with few reports from the UK (10-12). ER, PR and HER2 molecular markers are used often as surrogates for the intrinsic subtypes of breast cancer defined by mRNA expression profiling (13) because, unlike genetic profiling, the markers have been measured routinely in recent years. In the age of precision medicine, quantifying and monitoring cancer incidence by molecular subtypes is important in optimising public health prevention programmes, the allocation of resources and availability of screening, diagnostic and therapeutic services, and for improving outcomes (14). An important aspect of assessing trends by ER status is to account for missing data, as completeness of marker data has improved over time, but imputation methods can be applied to address this limitation (7-9, 15).

Within Scotland’s renowned, high-quality routine electronic health records, the Scottish cancer registry is an excellent resource to investigate temporal trends in cancer incidence. Data collection began for ER in 1997 and PR and HER2 in 2009 and so provides data almost a decade earlier than other UK national registries. While monitoring of breast cancer incidence in the UK is standard (16, 17), these data have not been presented by molecular subtypes. Further, recent changes in risk factors, such as, changes over time in reproductive factors and increasing obesity prevalence and alcohol consumption may have greater impact on ER+ specific breast cancer incidence rates compared to ER-given weaker/null associations with these subtypes (6, 18-20).

Here we report on breast cancer incidence trends in Scotland by ER and ER/HER2 combinations using several statistical methods: 1) Age-standardised and age-specific incidence rates, which are typically used to report cancer statistics (21); 2) Joinpoint regression models to determine whether significant changes occurred during 1997-2016, and the speed at which they have occurred (22); and 3) Age-Period-Cohort (APC) models (23-25) based on generalized linear model theory to enable description of age, period and birth cohort effects to provide possible clues to potential underlying factors contributing to incidence trends, and to thereby inform public health and NHS programmes.

## Methods

### Data and cohort definition

All primary invasive breast cancers (defined on the basis of the International Classification of Diseases, 10^th^ revision code of C50) diagnosed in women aged 20+ years, between 1997 and 2016 were ascertained from the Scottish cancer registry held by Information Services Division (ISD) of NHS National Services Scotland. The Scottish cancer registry achieves 98% breast cancer case ascertainment and is over 99% complete (26). **Supplemental Figure 1** describes the creation of the final study population. Women with primary breast cancer are the basis for analysis, each characterised by her worst-prognosis tumour. Permission for use of the data was obtained from the Public Benefit and Privacy Panel (PBPP) of NHS Scotland (reference number 1718-0057) and analyses were conducted in the Scottish National Safe Haven.

Additional demographic and tumour data obtained were: age at diagnosis, NHS Scotland regions (North, South East and West), tumour grade (grade I-well differentiated to III-poorly differentiated), tumour size (less than 10mm, 10-20mm and more than 20mm), nodal involvement (yes or no), screen-detected tumour (yes or no) and the status of molecular markers ER, PR and HER2 (positive, negative or unknown). ER and PR status is measured using immunohistochemistry (IHC) and HER2 status was assessed using fluorescent in-situ hybridization. Previous studies have noted that assessment of ER status reliability is high with an error rate below 5% (27). ER/HER2 combinations were used as surrogates for the four intrinsic subtypes of breast cancer, the gold standard for which uses mRNA expression profiling. ER+/HER2- was used as surrogate for Luminal A tumours, ER+/HER2+ for Luminal B, ER-/HER2+ for HER2 enriched tumours and ER-HER2- for triple negative tumours. The high quality of these data has been previously described (28).

### Statistical methods

Missing ER and ER/HER2 status were imputed conditioned on age and year of diagnosis, with the assumption that data were missing at random, using a validated method (7-9). Age-standardised incidence rates (ASR) per 100,000 women were calculated using the direct method, the European standard population (2013) (29) and mid-year estimates of the Scottish population for each age and year (30). Age-specific incidence rates were calculated for 5-year age groups (20-24 to 90+) and individual calendar years using two approaches: with number of tumours as the numerator for consistency with routine reporting and with one tumour per women as the numerator for all other analyses. ASR were calculated for all age groups combined and for three separate age-groups, with the middle group defined on the basis of eligibility for routine breast screening in Scotland, (20 to 49 years, 50 to 69 years and more than 70 years); and for each ER status and ER/HER2 combinations.

Joinpoint regression models were used to describe breast cancer incidence rates overall, by ER status and ER/PR/HER2 combinations for all women in the cohort and for three age groups (20-49, 50-69 and 70+ years). Joinpoint models describe if changes in the incidence trends occur and identify the time points at which a change is observed (referred to as joinpoints). The permutation test method, as described by Kim et al (22), was used iteratively: it starts by testing the null hypothesis of a simple model with zero joinpoints against the alternative hypothesis of a more complex model with the maximum number of joinpoints previously specified (3 joinpoints for this study). The procedure continues until all possible numbers of joinpoints have been tested. In the final model, the estimated annual percentage change (EAPC) for each of the periods identified is calculated. The average annual percent change (AAPC) is also reported as a measure of the overall trend from 1997 to 2016. Joinpoint regression software was used for all the analysis (31).

APC models were fitted for age-standardised incidence of ER+ and ER- tumours. The APC model provides a unique set of best-fitting log incidence rates obtained by maximum likelihood estimators for period, age and cohort, which has been shown to provide similar rates to ASR, but allows investigation of differences by birth cohorts--with the middle cohort as referent--which are not investigated in ASR or joinpoint regression analysis. As a consequence of small numbers in some strata, we restricted these models to women aged 30 to 85 years and used 28 two-year age groups (from 30-31 to 84-85) and 10 two-year periods (from 1997-1998 to 2015-2016) of calendar year of diagnosis, which covered birth cohorts from 1912 to 1986. The net drift, similar to the EAPC and AAPC estimate, is reported with 95% confidence intervals (CI). Local drifts were also estimated and describe the annual percentage change for each age-specific rate over time (32). In addition, period and cohort rate ratios are also presented to compare the age-specific rates in each period or cohort with the reference points in the middle of the study period and distribution of the birth cohort (2006 for period and 1949 for cohort). Along with cohort rate ratios, a combination test of significance for the complete cohort deviations is reported. This new combination test (32) aims to determine if there is an association of the observed rates with the birth cohorts above the linear influences represented by the net drift. The test provides a more robust method than the traditional Wald test while correcting for multiple testing. All statistical tests were two-sided and deemed statistically significant at the five percent level (p<0.05). With the exception of joinpoint regression, performed using specific software, all analyses were carried out using R (33).

Cumulative epidemiologic data supports a causal relationship between menopausal hormone therapy (MHT) and breast cancer risk (34, 35). To determine the possible proportion of women exposed, data from the Prescribing Information System in Scotland, which includes all items dispensed in Scotland was obtained from eDRIS to determine the number of MHT prescriptions and estimated proportions of prescriptions of women who might be exposed from 1996-2016.

## Results

### Characteristics of cohort by ER status

Between 1997 and 2016, 72,217 women of 20 years of age or older were diagnosed with at least one invasive breast cancer in Scotland (**Table 1**). Seventy six percent of these tumours were ER+, 16% were ER- and 8% had unknown ER status. However, the percentage of missing ER status decreased over time from 20% in 1997 to 2% in 2016. Proportions with unknown ER status differed by region and age: was higher in the West compared to the North and Southeast of Scotland and in women aged 70 years or older compared to women aged less than 70 years (14% missing vs 5%). Almost half of breast cancers were diagnosed among women of 50 to 69 years of age, the range for eligibility for routine breast cancer screening.

**Table 1:** Descriptive characteristics by ER status for all women with an invasive breast cancer diagnosed between 1997 and 2016 in Scotland.

| Characteristics |  | ER- |  | ER+ |  | ER unknown |  |
| --- | --- | --- | --- | --- | --- | --- | --- |
|  |  | n | % | n | % | n | % |
|  |  | 11726 | [16] | 55144 | [76] | 5347 | [8] |
| **Age at diagnosis** | <50 years | 3196 | (27) | 10550 | (19) | 695 | (13) |
|  | 50 to 69 years | 5668 | (48) | 28441 | (52) | 1580 | (30) |
|  | 70 years or older | 2862 | (24) | 16153 | (29) | 3072 | (57) |
| **Grade** | I-Well differentiated | 195 | (2) | 8288 | (15) | 232 | (4) |
|  | II- Moderately differentiated | 1714 | (15) | 25734 | (47) | 602 | (11) |
|  | III- Poorly differentiated | 8308 | (71) | 14586 | (26) | 642 | (12) |
|  | Unknown | 1509 | (13) | 6536 | (12) | 3871 | (72) |
| **Nodal Status** | Uninvolved/negative | 6194 | (53) | 29400 | (53) | 869 | (16) |
|  | Involved/positive | 4110 | (35) | 17369 | (31) | 415 | (8) |
|  | Unknown | 1422 | (12) | 8375 | (15) | 4063 | (76) |
| **Screen detected** | No | 9622 | (82) | 37400 | (68) | 4047 | (76) |
|  | Yes | 1943 | (17) | 17119 | (31) | 490 | (9) |
|  | Unknown | 161 | (1) | 625 | (1) | 810 | (15) |
| **Tumour size** | Less than 10mm | 1017 | (9) | 6470 | (12) | 202 | (4) |
|  | 10 to 20 mm | 3428 | (29) | 20449 | (37) | 478 | (9) |
|  | More than 20mm | 4960 | (42) | 18168 | (33) | 512 | (10) |
|  | Unknown | 2321 | (20) | 10057 | (18) | 4155 | (78) |
| **PR status\*** | Negative | 3803 | (79) | 3036 | (12) | <10 | (<1) |
|  | Positive | 226 | (5) | 15869 | (62) | <10 | (<1) |
|  | Unknown | 764 | (16) | 6489 | (26) | 901 | (99) |
| **HER2 status\*** | Negative | 2761 | (66) | 18709 | (84) | 36 | (5) |
|  | Positive | 1210 | (29) | 2553 | (11) | 10 | (1) |
|  | Unknown | 184 | (4) | 1129 | (5) | 725 | (94) |

Brackets [ ] indicate row percentages and parenthesis ( ) indicate column percentages for that category. \*Denotes markers that were recorded from 2009 to 2016, and number of cases for those years=31,099. Differences by known ER status for all characteristics were significantly different with $\chi^2$ $p<0.001$ .

Tumour characteristics differed by ER status, with ER- tumours having characteristics associated with more advanced/aggressive disease. ER- tumours had higher grade, were larger, more likely to have positive lymph node status and less likely to be screen detected than ER+ tumours. The patterns of other molecular markers also differed by ER status, with ER- tumours more likely to be PR- and HER2+ than ER+ tumours. In contrast, ER+ tumours were more likely to be PR+ and HER2- than ER- tumours.

The combinations of ER/HER2 status **Figure 1a** show that among women with known ER/HER2 status (N=27,580) most tumours were ER+/HER2-, with ER-/HER2+ tumours being the least common combination. ER-/HER2- tumours, the most aggressive subtype, was the second most common at 11%. Cross-sectional age specific curves for the ER/HER2 combinations (**Figure 1b**) show incidence of all subtypes increasing rapidly with age until the approximate age of menopause, age 50 years, after which the increase continues more gradually up to 70 years for ER+/HER2- tumours but did not increase further for ER- tumours or ER+/HER2+ tumours.

**Figure 1:**
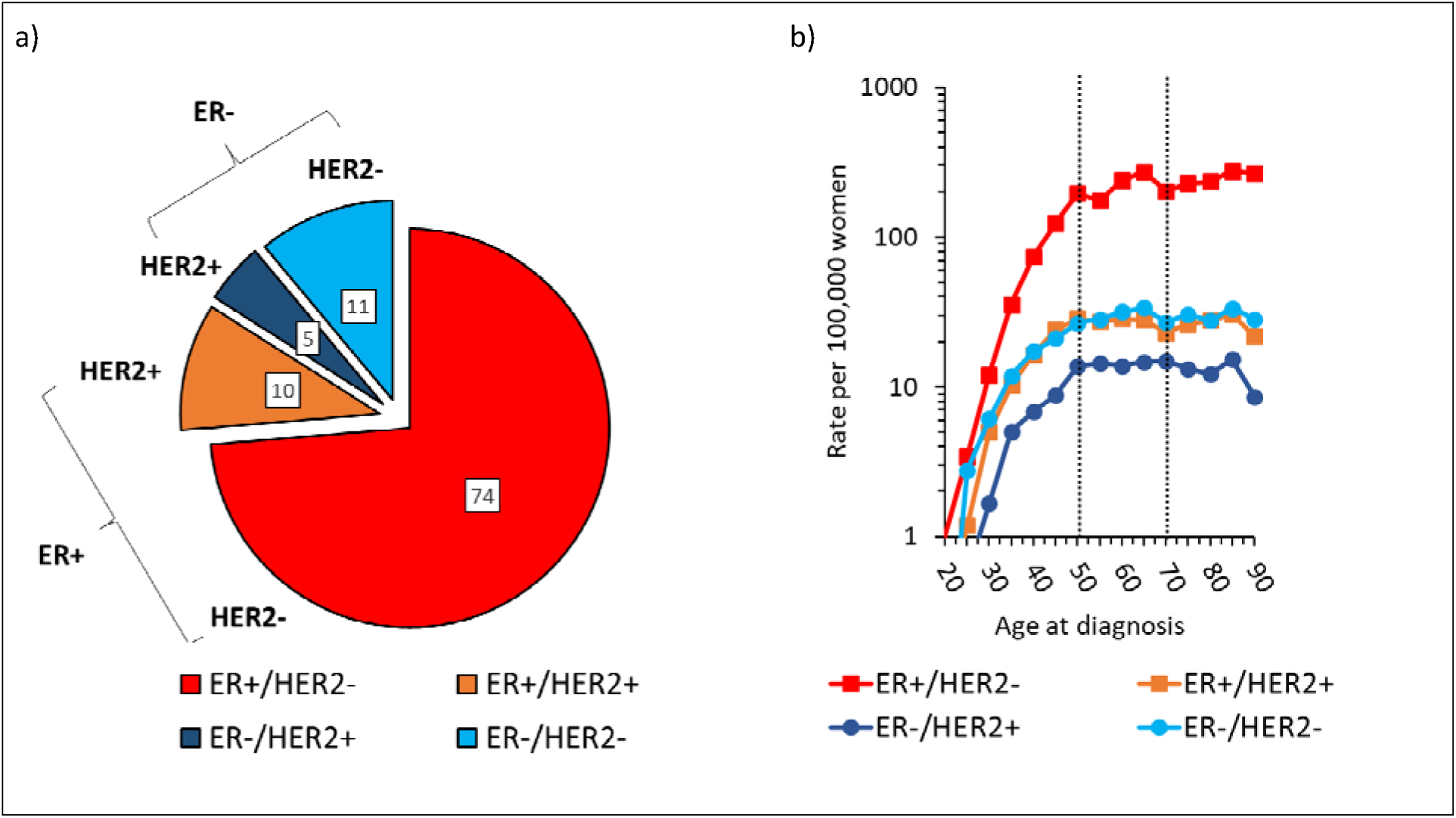
Distribution of the breast cancer subtypes by ER/HER2 status and their age-specific incidence in Scotland for 2009-2016(N=31,099) Panel (a) shows a pie chart and panel b) shows age specific incidence on the log scale by subtype (b) Data are for 31,099 breast cancer cases with ER/HER2 status recorded and missing status imputed for analysis. Dotted lines in graph denote ages 50 to 70 years, the age-group invited for screening in Scotland every 3-years.

### Age standardised incidence rates with EAPCs from joinpoint regression

As would be expected, increases in breast cancer incidence over time are less marked when using worst tumour characteristics per person (so that subjects are only counted once) rather than tumours as the numerator for incidence. Age-standardised incidence of ER+ tumours increased from 98 per 100,000 women in 1997 to 113 per 100,000 women in 2016 (**Figure 2**), with an average annual percentage change (AAPC) of 0.4% (95% CI: −0.1 to 1%). Incidence of breast cancer was higher for ER+/HER2- tumours than for the rest of the subtypes (**Figure 2**) and the time trend for incidence this subtype was similar to that of ER+ tumours with increases observed up to 2011. ER+/HER2+ and ER-/HER2+ tumour incidence decreased over time and there appeared to be an increase ER-/HER2- tumour incidence from 2011, although the latter finding was based on relatively small numbers.

**Figure 2:**
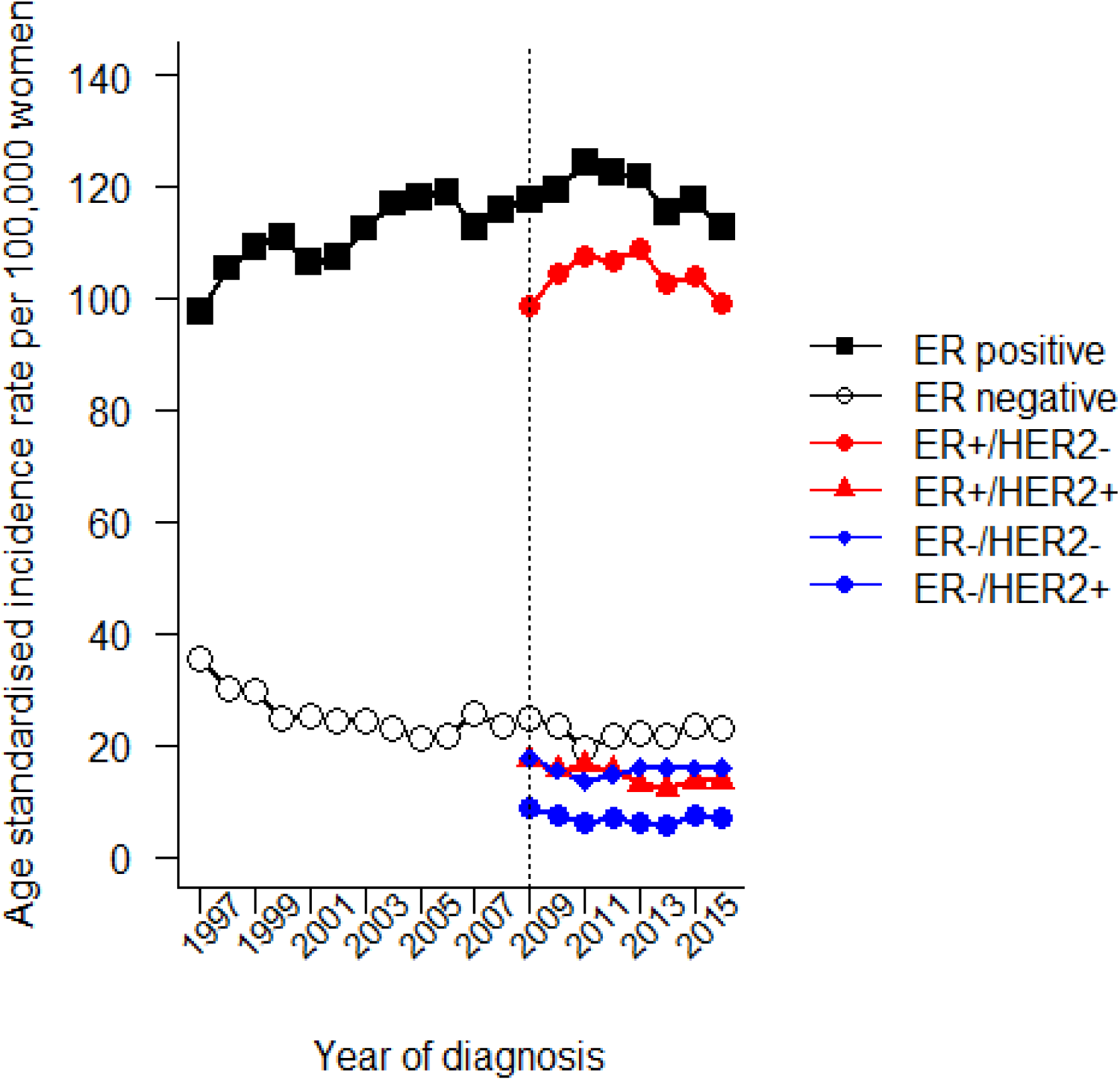
Age-standardised breast cancer incidence rates by ER and HER2 status in Scotland for 1997-2016. ER status was available for the entire period and data are for N=72,217 breast cancer cases. ER/HER2 combinations were available from 2009 to 2016, N=31,099. Dotted line at year 2009 denotes when HER2 status started to be collected in the Scottish cancer registry. Rates are age-standardised to the 2013 European population and missing ER and HER2 marker status were imputed based on age and year of diagnosis (see methods).

Estimates from the joinpoint analysis (**Table 2**) show that the increase in incidence of ER+ tumours was reasonably constant (1.2% increase annually, 95% CI: 0.8 to 1.5%) from 1997 till around 2012 after which incidence dereased by approximately 2.2% annually (95% CI: - 4.7 to 0.4%). By contrast, ER- tumour incidence decreased consistently over the study period by approximately 2.5% per year (95% CI: −3.9 to −1.1%). Joinpoint regression results by age-group in **Table 2** revealed that women of 50 to 69 years of age had the highest increases in ER+ incidence at a similar period as noted overall (**Figure 3a**); followed by women aged 20 to 49 years where ER+ tumour incidence increased by 1.1% annually. For older women of 70 years or more rates were stable. The decreases observed in ER- tumours were consistent across the three age groups (**Figure 3b**).

**Table 2:**
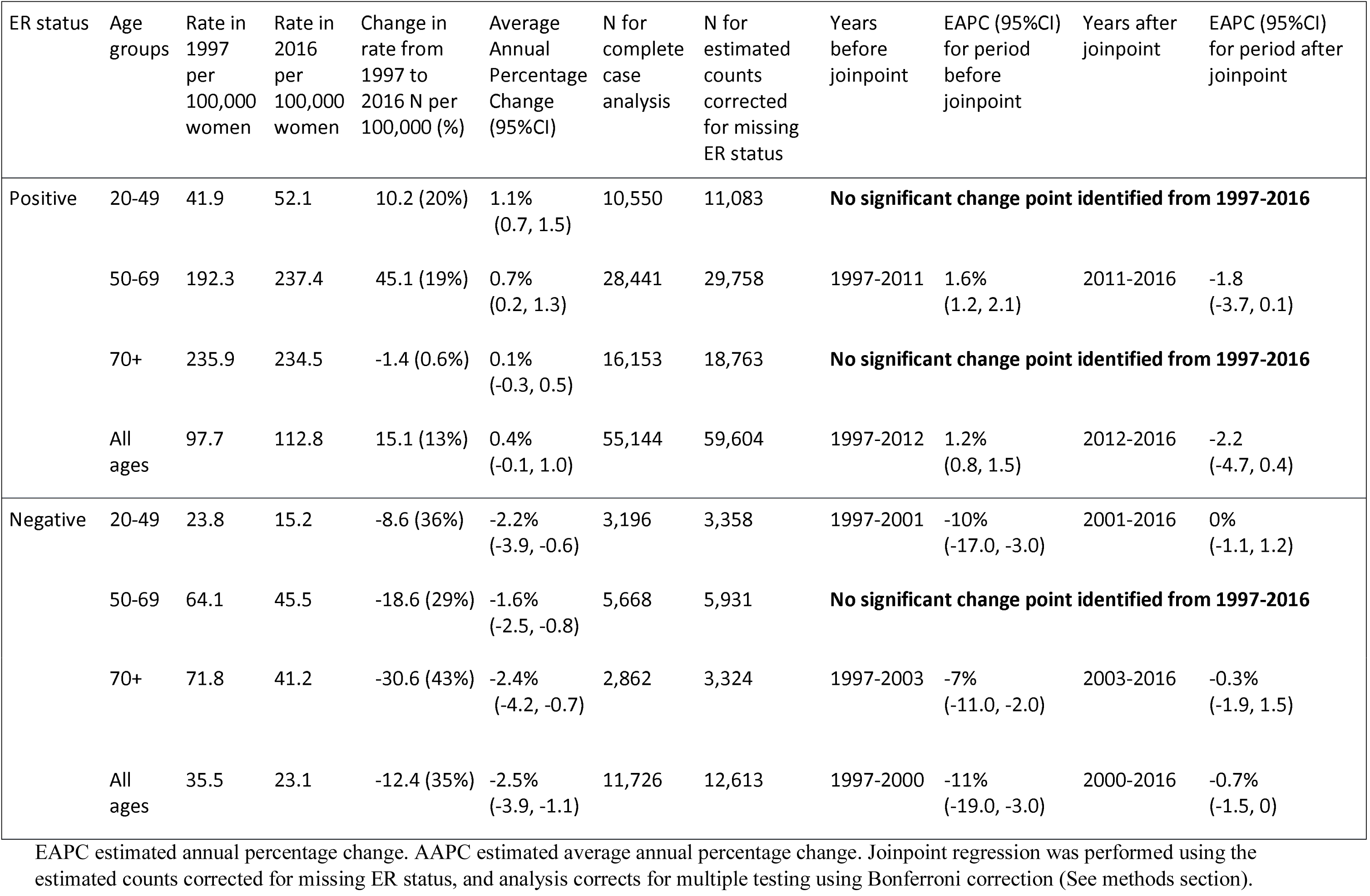
Joinpoint regression analysis stratified by age groups and ER status from 1997 to 2016.

**Figure 3:**
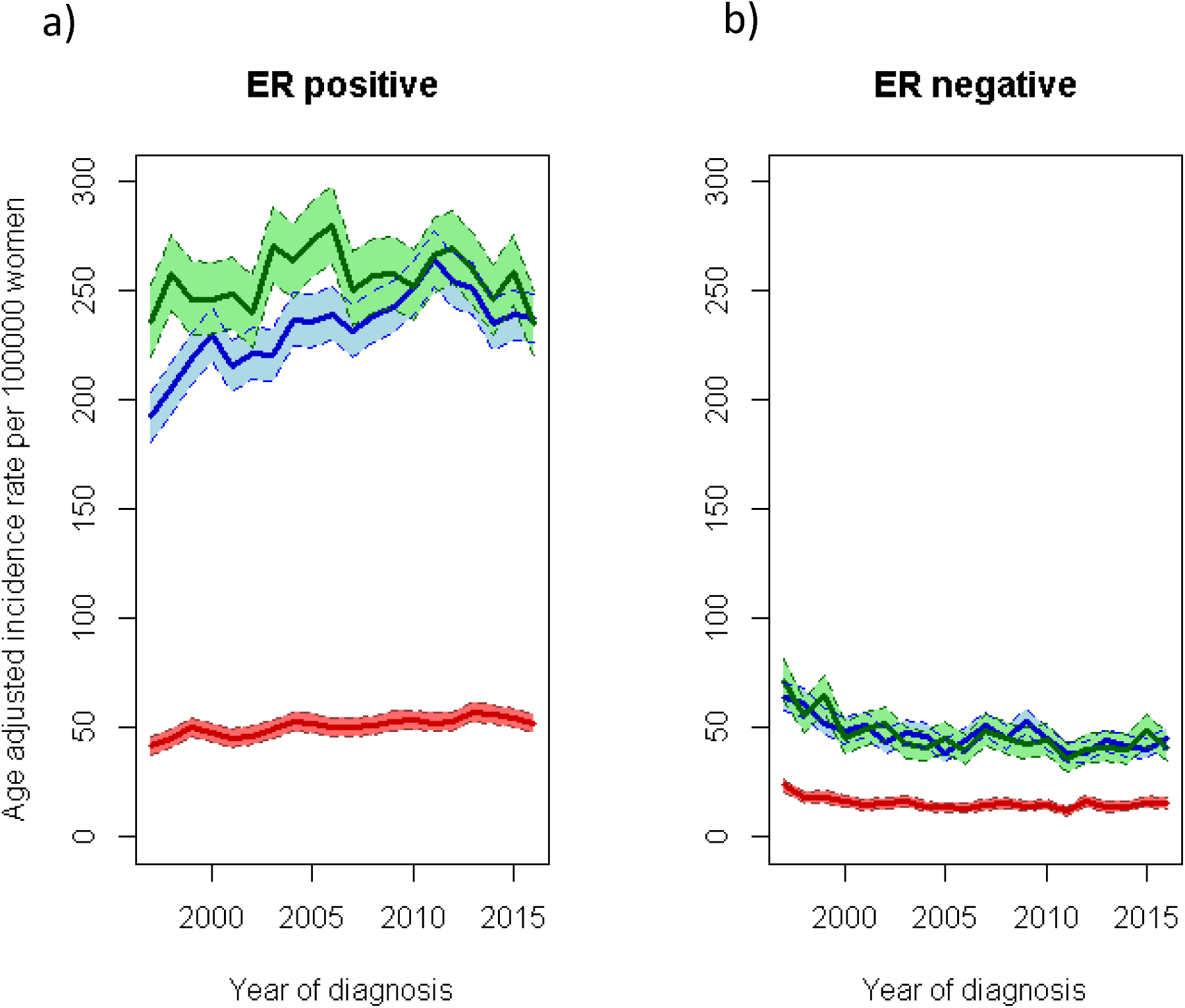
Age-specific trends in breast cancer incidence stratified by age groups and ER status in Scotland for 1997-2016. ER positive (a) and ER negative (b) age-specific trends for age groups 20-49 (red), 50 to 69 (blue) and 70 years old (green). Shaded areas surrounding lines indicate 95% CI of rates.

Additional joinpoint regression analysis by the ER/HER2 combinations of breast cancer (**Supplemental Table 1**), revealed that ER+/HER2- and ER-/HER2+ breast cancer incidence remained constant from 2009 to 2016. In contrast, ER+/HER2+ incidence decreased for all women, and that decrease was predominantly among women aged more than 50 years old. ER-/HER2- breast cancer incidence slightly increased from 2011 to 2016 and was most marked among women aged 20 to 49 years.

### Age-Period-Cohort models

Results from APC models were consistent with those observed from joinpoint regression, with net drifts suggesting increases in the overall incidence of ER+ tumours by 0.8% per year (95% CI: 0.6 to 1.0%/year) from 1997 to 2016, and ER- tumour incidence decreasing by 1.4% (95% CI: −1.8 to −1.1%/year). After adjusting for period and cohort effects, local drifts showed that the highest increase in incidence of ER+ tumours was observed in women around 70 years of age (2% per year, 95%CI: 1.6 to 2.4%) (**Supplemental Figure 2a**). The greatest drop in incidence of ER- tumours was observed in women between 53 and 61 years of age (**Supplemental Figure 2b**).

Compared to the women born in 1949, ER+ tumour incidence was higher among more recent birth cohorts. In contrast, ER- incidence was lower for more recent birth cohorts compared to the cohort born in 1949. Cohort rate ratios (CRRs) compared to women born in 1949 ranged from 0.7 for women born in 1913 to 1.8 for women born in 1985 for ER+ tumours and from 1.5 for women born in 1913 to 0.5 for women born in 1985 for ER- tumours (**Figure 4**). The combination test for ER+ tumours revealed cohort effects beyond the log-linear trend shown by the net drift (p value=6.3 × 10^−8^) but the test for ER- tumours failed to reach significance (p value=0.14).

**Figure 4:**
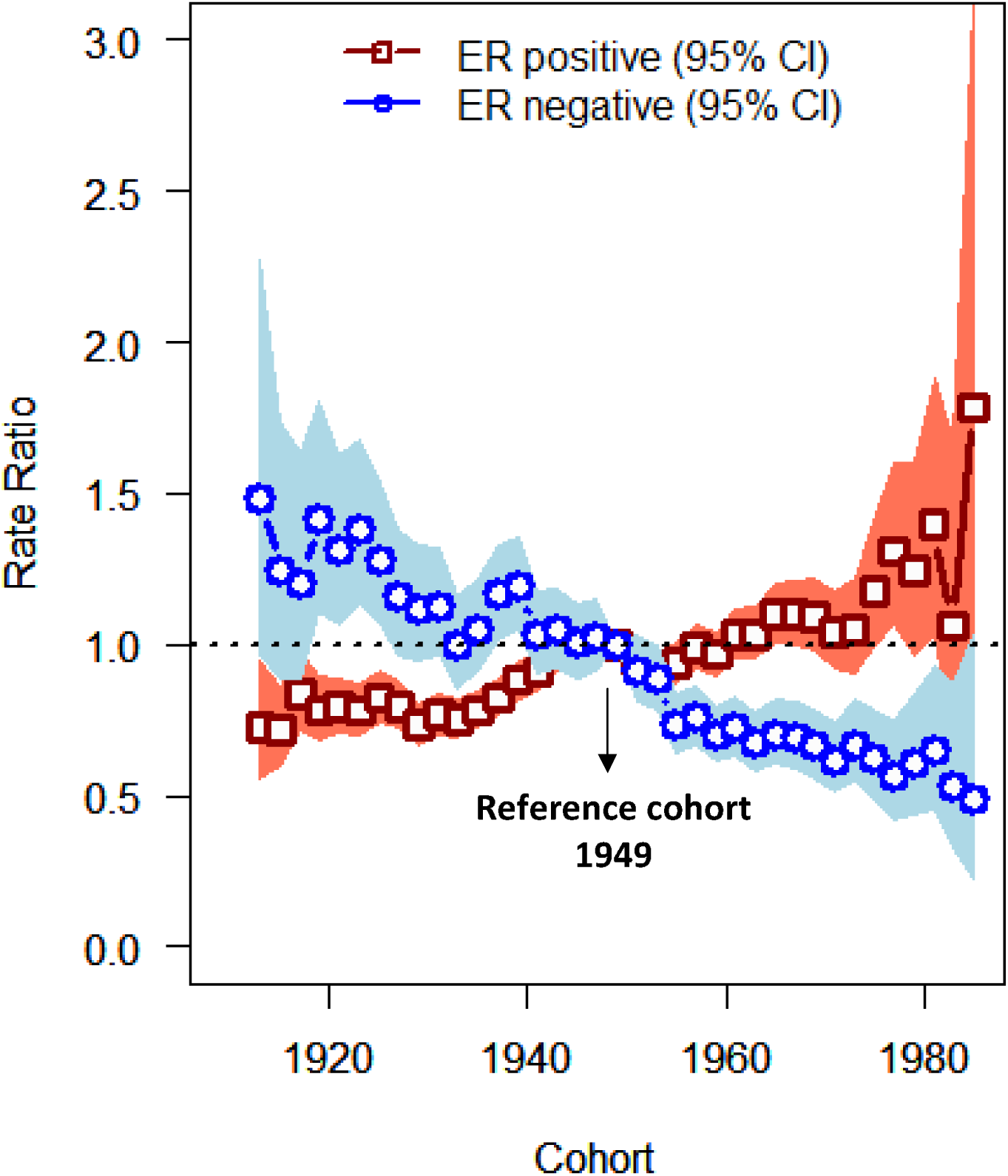
Birth cohort rate ratios (CRR) for breast cancer incidence rates in Scotland by ER status. CRR describe the incidence rates for each birth cohort relative to the 1949 birth cohort

### MHT use in Scotland

**Supplemental Table 2** shows that MHT utilisation has changed dramatically since 2003 in Scotland and use steadily declined following the results of the Women’s Health Initiative trial indicating increased risks of cardiovascular disease and breast cancer among women randomised to MHT (36). Data from the Prescribing Information System in Scotland, which includes all items dispensed in Scotland, show that numbers of MHT-prescriptions and estimated proportions of women who might be MHT-exposed at 40-79 years of age as high as 16.5% in 1999 but as low as 4.7% by 2015.

## Discussion

This study demonstrates that In Scotland, temporal trends of breast cancer incidence were distinct by molecular subtypes, with increases for ER+ and decreases for ER- tumours between 1997 and 2016. With respect to ER+ tumours, their incidence increased for all ages for the study period but particularly among women of screening ages 50 to 69 years, with the largest increases occurring from around 1997-2011 followed by modest declines. In contrast, the incidence of ER- cancers decreased among all ages till the early 2000’s. Finally we noted cohort effects such that in comparison to women born around 1950, women of older generations (those born in the 1910s to 1940s) had a lower risk of ER+ tumours, whereas there was no significant evidence for cohort effects for ER- tumours. Further analysis of the incidence trends by subtype (as defined by ER/HER2 combinations) showed similar results to those observed by ER status only. ER+/HER2- (surrogate for luminal A) tumours followed the same pattern as ER+ tumours, and ER-/HER2- (surrogate for Triple Negative) similar to ER- tumours.

Consistent with other reports from the United States, Denmark and Ireland (7-9), our data show for the first time in a UK national cancer registry, contrasting temporal trends of breast cancer incidence by ER status and suggest the presence of aetiologic heterogeneity with distinct patterns by period, age at diagnosis and birth cohort. Previous studies have shown estimated annual increases in the age-standardised rate of breast cancer from early 90’s to the 2010 for ER+ ranging from 0.1 to 3% and declines for ER- ranging from −1.9 to −3.4% (7-9). The Scottish Cancer Registry’s detailed tumour hormone receptor data have been used to describe trends in incidence patterns of breast cancer. Specifically, it was previously reported that there were declines in ER positive tumours among women 50–64 years of age that was statistically significant by 2005 (12). These findings were attributed to reduction in MHT use, which had been shown to be associated with increased risk of breast cancer (34, 36). Unlike the previous analysis we excluded women with a previous malignancy, imputed missing ER status, used individuals rather than tumours as the numerator for incidence rates but the findings were similar for comparable years. The declines in breast cancer incidence observed in Scottish data have been also shown in the United States (7, 37-39), Sweden (8) and France (40). We observed consistent increases over time for ER+ tumour incidence beyond 2002 after which MHT use declined. Based on recent reports on the association of MHT use and breast cancer risk (35) MHT has been estimated to contribute to 1 in 20 breast cancers diagnosed worldwide from 1990. In more recent years when MHT use has declined in use, it has been estimated to have an approximate 5 year lag time to breast cancer incidence and contribute to 2.3% of breast cancers in Scotland in recent years (41). Despite reductions in HRT use between 2005-2011 incidence of breast cancer continued to increase. In addition to long-term effects of previous HRT use, other factors, such as screening, obesity and alcohol consumption (41) are likely also to contribute to time trends in breast cancer incidence.

Mammographic screening is likely to be an important contributing factor to the increased incidence of ER+ tumours we observed from 1997-2011. In Scotland, the breast screening programme was established in 1988 with full national coverage attained in 1991 (42). Scotland’s breast screening programme was introduced earlier than in other countries that have evaluated breast cancer incidence trends by ER status (ie. 2000 in Ireland and 2010 in Denmark, in the US there are no national screening programmes). In Scotland, from 1994-2003 women of 50-64 years of age were invited for screening, which was extended to include women of 65 to 70 years of age in 2003. Over the course of the entire study period in Scotland, the mammographic screening programme had around 75% uptake. Our data showing ER+ tumours are more likely to be screen detected than ER- tumours (31% vs 17%) and our APC model results showing incidence of ER+ tumours greatest for those of screening ages between 65 and 72 years, suggest that some of the increases observed in ER+ tumours is likely to be due to detection of prevalent disease in these older women. A similar pattern was also observed in the previous report (12). In contrast, screening is probably not a major contributor to the more recent declines in incidence that we observed for ER+ tumours starting in 2011 among women aged 50 to 69 years. Some studies have suggested that decreases in incidence may coincide with the saturation of the screening programme (7, 38), however, given that uptake in Scotland remained approximately stable for the whole study period with a slight fall (0.6%) for the last three-year period (42) this seems unlikely to be a contributing factor.

Obesity prevalence and alcohol intake have increased over time, although declines for alcohol in particular have been noted since 2003. Recent analysis estimates these two factors account for 18% of all breast cancers diagnosed in Scotland (41). The relationships of obesity and alcohol use are complicated and seem to be modified by menopausal status, but previous analysis suggest increased alcohol use to be associated with increased risk of ER+ tumours. Obesity in premenopausal women is particularly associated with a higher risk of ER-negative compared with ER-positive cancer (43-45). Secular trends in increasing obesity prevalence may contribute to the divergent trends in breast cancer incidence by ER status observed in younger women. Our analysis although showing overall increases in incidence for ER+ breast cancers for the entire study period, suggests for the first-time incidence of ER+ tumours not rising or slightly declining, particularly for women aged 50 to 69 years of age that started around 2011. This recent decrease in ER+ tumours incidence parallels results in Ireland showing incidence of ER+ tumours stabilizing or slightly decreasing from 2008 to 2013 (9). From national survey data in Scotland, obesity prevalence increased between 1995 and 2008 and then remained approximately stable until 2012 after which there was some evidence of declines especially among the most affluent groups of women (46). Future work is required to clarify the contribution of changes in obesity prevalence to trends in incidence of breast cancer and the lag time in its contribution to breast cancer incidence, including short and long term effects.

In addition to obesity, other factors seem to have a differential effect on risk of ER+ and ER- tumours. For example, reproductive factors such as younger age at menarche, older age at first birth, nulliparity, lack of breastfeeding and older age at menopause are associated with an increased risk of ER+ breast cancers (3, 47). Parous women have an increased risk of triple negative or basal-like tumours compared to nulliparous women (48-50). Breastfeeding has been consistently associated with a reduced risk particularly more aggressive subtypes of breast cancer, namely ER-(48) and triple negative tumours (47, 48). In Scotland, reproductive patterns have changed considerably over time: the average age of a woman at first birth has increased from 26 years in 1975 to 30.5 years in 2017, fertility rates have decreased and the number and proportion of women not having children has also increased in recent decades (51, 52). Our study provides evidence of changes in incidence of breast cancer between birth cohorts by ER status with higher year of birth positively associated with risk of ER+ breast cancer and inversely associated with risk of ER-. The reproductive factor changes that have occurred in Scotland may be associated with the increasing ER+ breast cancer incidence in more recent birth cohorts. Breastfeeding practice at 6-8 weeks has increased in Scotland from 36% in 2001 to 42% in 2018 (53), and this may have also contributed to some of the declines observed for ER- tumours. Current, estimates suggest 5.2% of breast cancer attributable to not breastfeeding in Scotland (41).

The strengths of our study are the high quality of the longitudinal data collected within the Scottish cancer registry, the first one in the UK and fifth in the world that routinely started recording molecular marker data (ER status from 1997 and PR and HER2 status from 2009). Marker data can be used to monitor and describe incidence trends in the future and for other types of cancer that also display heterogeneity. Further, monitoring breast cancer incidence by molecular subtypes can help the NHS allocate resources for treatment and prevention and lead to the identification of high-risk groups of women for which to implement future prevention programmes and treatments.

Limitations of our study include the absence of individual level risk factor data. However, future studies should be possible to identify some key factors using linked data. Scotland is renowned for its high-quality longitudinal data and ability to perform linkage studies using a unique identifier. Hence, we envision future analysis using the Scottish cancer registry linked to other datasets, including community prescription drug records, maternity records and hospital records to provide more detailed information on the role of patterns of key risk factors in breast cancer incidence trends. Another limitation of the study is the lack of mRNA expression assays for the classification of the molecular subtypes of breast cancer. In our study, markers measured by IHC are used as surrogates for the molecular subtypes, which are reasonably good proxies but mRNA profiling data would be considered a gold-standard for intrinsic-subtype classification (54).

In conclusion, incidence trends of breast cancer in Scotland differ by ER status and are consistent with trends observed in other countries. Additional data are needed to establish whether HER2+ tumours, which are ER-, remain low since trastuzumab is one of the costliest breast cancer treatments used by the NHS. Further research should be focused on monitoring incidence trends by subtype given notable risk and treatment differences for breast cancer subtypes.

## Data Availability

As these are NHS patient data, approval to obtain individual data are needed. Summary estimates and counts where they would reduce identifiability are presented.

## Additional information

### Ethics approval and data Availability

The data used in this study can be accessed through application to electronic Data Research and Innovation Service (eDRIS), a part of the Information Services Division of NHS Scotland. Approval from the Public Benefit and Privacy Panel for Health (PBPP) and Social Care is a requirement for data access. Our project was approved by PBPP reference number 1718-0057.

### Authors’ contributions

Conception & Design of the study: JDF, SW, IME, SB

Interpretation of data: All authors

Drafting of the manuscript: JDF, IME, SW

Revised work and provided important intellectual content: All authors

Final approval of the manuscript: All authors

## Acknowledgements

We thank NHS Scotland and Information Services Division for collection of data used for this analysis. We specifically would like to acknowledge Andrew Deas, Suhail Iqbal, Rita Nogueira and Ross Murdoch of NHS National Services Scotland.

